# Effectiveness of one dose of killed oral cholera vaccine in an endemic community in the Democratic Republic of the Congo: A matched case-control study

**DOI:** 10.1101/2023.08.07.23293369

**Authors:** Espoir Bwenge Malembaka, Patrick Musole Bugeme, Chloe Hutchins, Hanmeng Xu, Juan Dent Hulse, Maya N. Demby, Karin Gallandat, Jaime Mufitini Saidi, Baron Bashige Rumedeka, Moïse Itongwa, Esperance Tshiwedi-Tsilabia, Faida Kitoga, Tavia Bodisa-Matamu, Hugo Kavunga-Membo, Justin Bengehya, Jean-Claude Kulondwa, Amanda K Debes, Nagède Taty, Elizabeth C. Lee, Octavie Lunguya, Justin Lessler, Daniel T Leung, Oliver Cumming, Placide Welo Okitayemba, Daniel Mukadi-Bamuleka, Jackie Knee, Andrew S Azman

**Affiliations:** Department of Epidemiology, Johns Hopkins University, Baltimore, MD 21205, USA; Centre for Tropical Diseases and Global Health (CTDGH), Université Catholique de Bukavu, Bukavu, Democratic Republic of the Congo; Department of Disease Control, London School of Hygiene & Tropical Medicine, London, UK; Ministère de la Santé Publique, Zone de Santé d’Uvira, Democratic Republic of Congo; Rodolphe Merieux INRB-Goma Laboratory, Goma, North-Kivu, Democratic Republic of the Congo; Institut National de Recherche Biomédicale, INRB, Kinshasa, Democratic Republic of the Congo; Ministère de la Santé Publique, Hygiène et Prévention, Division Provinciale de la Santé Publique du Sud-Kivu, Democratic Republic of the Congo; Department of International Health, Johns Hopkins University, Baltimore, MD 21205, USA; PNECHOL-MD, Community IMCI, Ministry of Health, Democratic Republic of the Congo; Service of Microbiology, Department of Medical Biology, University of Kinshasa, Kinshasa, Democratic Republic of the Congo; University of North Carolina Population Center, University of North Carolina at Chapel Hill, Chapel Hill, NC, USA; Department of Epidemiology, Gillings School of Global Public Health, University of North Carolina at Chapel Hill, Chapel Hill, NC, USA; Division of Infectious Diseases, University of Utah School of Medicine, Salt Lake City, Utah, USA; Division of Microbiology and Immunology, University of Utah School of Medicine, Salt Lake City, Utah, USA; Geneva Centre for Emerging Viral Diseases, Geneva University Hospitals, Geneva, Switzerland; Division of Tropical and Humanitarian Medicine, Geneva University Hospitals, Geneva, Switzerland

## Abstract

**Background:** A global shortage of cholera vaccines has increased the use of single-dose regimens, rather than the standard two-dose regimen. There is limited evidence on single-dose protection, particularly in children. In 2020, a mass vaccination campaign resulting in largely single dose coverage, was conducted in Uvira, an endemic urban setting in eastern Democratic Republic of the Congo. We examined the effectiveness of a single-dose of the oral cholera vaccine Euvichol-Plus^®^ in this high-burden setting.

**Methods:** We recruited medically attended confirmed cholera cases and age-, sex-, and neighborhood-matched community controls during two distinct periods after mass vaccination, October 2021 to March 2022 (12–17 months post-vaccination) and October 2022 to October 2023 (24–36 months post-vaccination). The odds of vaccination in cases and controls were contrasted in conditional logistic regression models to estimate unadjusted and adjusted vaccine effectiveness.

**Findings:** We enrolled 658 confirmed cases and 2,274 matched controls during the two study periods with 15·0% of cases being under five years old at the time of vaccination. The adjusted single-dose VE was 52·7% (95% CI: 31·4–67·4) 12–17 months post-vaccination and 45·5% (95% CI: 25·8– 60·0) 24–36 months post-vaccination. While protection in the first 12–17 months post-vaccination was similar for 1–4-year-olds and older individuals, over the third year post-vaccination the estimate of protection in 1–4 year-olds (adjusted VE 33·1%; 95% CI: -30·0–65·6) appeared to wane with confidence intervals spanning the null.

**Interpretation:** A single-dose of Euvichol-Plus^®^ provided substantial protection against medically attended cholera for at least 36 months post-vaccination in this cholera endemic setting. While our evidence provides support for comparable levels of protection in young children and others in the short-term, protection among young children may wane significantly by the third year after vaccination.

**Funding:** Wellcome Trust and Gavi (GAVI-RFP-2019-062).

**Research in context:** *Evidence before this study:* In late 2022, due to increasing demand for killed, whole-cell, oral cholera vaccines (kOCV) and limited production capacity, the International Coordinating Group (the organization managing emergency stocks of kOCVs) changed policy to deploy single-dose, rather than the standard two-dose regimen, for emergency vaccination campaigns. This decision was in line with WHO guidance on the use of a single dose in outbreaks, where short-term protection is key. However, this recommendation is based on a limited number of clinical studies with short-term follow-up. There is also limited evidence on the magnitude and duration of protection conferred by a single dose of kOCV, particularly in children under five years of age. We searched PubMed for randomized trials and observational studies published in English before November 1, 2023, that reported estimates of protection conferred by a single dose of kOCV, using the term “(effectiveness OR efficacy) AND cholera* AND vaccine”. We found no published studies estimating the effectiveness of a single dose of Euvichol-Plus^®^, and only one study reporting two-dose effectiveness. Despite this paucity of evidence, this is the only vaccine currently available in the global stockpile. To date, there has been one randomized trial conducted in Bangladesh between 2014 and 2016, and seven observational studies conducted between 2009 and 2016 in Guinea, Haiti, India, Malawi, Sudan, Zambia and Zanzibar, reporting effectiveness estimates of a single dose of the current generation of kOCV. Aside from the trial in Bangladesh, all estimates were based on secondary analyses that the studies were not powered to estimate. The Bangladesh trial is the only study to date that provides an age-stratified estimate of single-dose protection, and while it found an overall protective efficacy of 62% (95% CI: 43– 75) during the 2-year follow up for individuals aged five years or older, it found no significant protection conferred by the Shanchol kOCV (a bioequivalent of Euvichol-Plus^®^) for individuals under five years of age (protective efficacy: -44%, 95% CI -220 to 35). Four of the seven observational studies provide single-dose vaccine effectiveness (VE) estimates only during the first 12 months post-vaccination with estimates ranging from 43% (95% CI -84-82) in Guinea to 93% (95% CI 69-98) in Haiti. The three other observational studies providing a single dose VE estimate between 12-30 months post-vaccination were unable to demonstrate statistically significant protection conferred by kOCV, with estimates ranging between 32·5% (95% CI - 318·0-89·1) in India and 40% (95% CI -31-73) in Haiti. No vaccine protection estimates have been published from the two identified cholera endemic foci in Africa, the Democratic Republic of the Congo and Nigeria.

*Added value of this study:* In this vaccine effectiveness study, we show that a single dose of Euvichol-Plus^®^ vaccine can provide significant protection against medically attended cholera for up to 36 months after vaccination in a cholera endemic setting in Africa, though protection in children under five years old remains unclear. These estimates help fill critical gaps in our understanding of the magnitude and duration of protection from a single dose of the most widely used kOCV, Euvichol-Plus^®^ and is one of only a few studies to measure protection in an endemic setting in Africa.

*Implications of all the available evidence:* The corpus of available evidence suggests that use of a single dose of kOCV in emergency situations where cholera is endemic, like Uvira, is justified and that providing a second dose within the first 12-24 months post-vaccination may only provide marginal benefit to the general population. However, more evidence and analyses are needed to weigh the costs and benefits of tailored vaccination approaches for those under five years old, including possibility of providing a second dose at an earlier timepoint.

## Introduction

Safe water, sanitation, and hygiene (WASH) is the cornerstone of cholera prevention and control. While universal access to safely-managed WASH services remains the ultimate priority, this is likely a distant prospect.^1^ Killed whole-cell oral cholera vaccine (kOCV) is an effective short-term intervention to reduce cholera risk in high-burden settings and is a key component of the global roadmap to end cholera.^2^ kOCVs are typically delivered as a two-dose regimen that provides protection for at least three years.^3,4^ In a meta-analysis of kOCV protection, estimated two-dose efficacy was 58% (95% CI: 42 to 69), over an average of 28 months post-vaccination. Lower protection was noted among young children.^3^

The Euvichol-Plus^®^ vaccine (Eubiologics, Seoul, Republic of Korea) is currently the only WHO-prequalified kOCV manufactured and included in the global stockpile after the Shanchol^®^ (Shantha Biotechnics, Hyderabad, India) ceased production in 2023.^5^ Euvichol-Plus^®^ is considered a bioequivalent of Shanchol^®^.^6^ Almost all evidence of kOCV clinical protection is based on the studies carried out on Shanchol^®^,^7–9^ although one observational study explored the protection conferred by a Euvichol-Plus^®^ two-dose regimen.^10^

Demand for kOCVs outstripped the global supply in 2022, with only 33 million doses distributed out of the 72 million requested.^11^ In late 2022, the International Coordinating Group, which manages the global emergency stockpile for cholera vaccines, suspended the provision of the standard two-dose regimen in emergency vaccination campaigns, replacing it with a single-dose regimen due to limited vaccine supply.^12^ However, there are limited data on the protection offered by one dose of kOCV over extended periods (>12 months) or among children 1-4 years old.

Only a few studies have estimated single-dose protection in the general population, with point estimates suggesting short-term protection up to 16 months after vaccination in Haiti, though with large uncertainty. ^7–9,13–16^ A randomized trial in Bangladesh, the only study to provide age-stratified estimates of single-dose protection, suggested that Shanchol^®^ conferred no protection in 1-4 year-olds in the first six months post-vaccination, despite significant protection in older individuals for at least two years.^17,18^

In late 2020, the Ministry of Health of the Democratic Republic of the Congo (DRC) conducted a mass vaccination campaign of Euvichol-Plus^®^ in the city of Uvira in the South Kivu province. The estimated coverage of the vaccination campaigns was low, and as most vaccinated individuals reported receiving only one dose, we assessed effectiveness of a single dose of kOCV during outbreaks that occurred 12-17 and 24-36 months after vaccination.

### Study design, setting and vaccination campaigns

We conducted a matched case-control study in Uvira, a city of approximately 280,000 inhabitants on the northwestern shore of Lake Tanganyika with sporadic armed conflict, socio-political instability, and population displacement. Cholera cases are detected year-round in Uvira since 1978, often with distinct seasonal peaks and notable historical outbreaks.^19,20^ Household surveys conducted in 2016/17 in Uvira indicated surface water as the main drinking water source for 37.2% of households,^21^ and in areas close to the rivers and with the lowest tap water availability, >80% of households use drinking water contaminated with *Escherichia coli*.^*22*^ The same surveys estimated that about half (48.2%) of the population relied exclusively on tap water for drinking needs ^21^, and a recent study showed that between 2017 and 2021, the water service quality remained suboptimal, or deteriorated in many parts of the city.^23 20^

In April 2020, severe flooding caused at least 54 deaths, the displacement of approximately 80,000 people, and substantial damage to housing and WASH infrastructure in Uvira, prompting the Ministry of Health to conduct emergency cholera vaccination campaigns.^24^ Vaccination took place in two rounds, from 29 July to 8 August and 28 September to 05 October 2020 targeting all individuals in Uvira one year and older. The campaigns included door-to-door vaccination for five days, followed by vaccination offered through health facilities. While two rounds of vaccination were implemented, in a representative household survey we conducted 11 months after vaccination, 23% (95% CI: 20-27%) of the participants reported receiving two doses of the vaccine and 32% (95% CI: 38-36%) reported receiving one dose. No kOCV has been administered in this population between these campaigns and the end of our study period, 18 October 2023.

### Clinical cholera surveillance system in Uvira

Our study is based on enhanced clinical surveillance of cholera implemented at the two official health facilities designated to treat cholera patients in Uvira, the cholera treatment centre at the Uvira General Referral Hospital and the cholera treatment unit at the Kalundu CEPAC health centre (henceforth, “CTCs”). We attempted to identify and recruit all patients at least 12 months old with acute watery diarrhoea within the 24 hours prior to the admission to the CTCs (referred to as ‘suspected cholera’). Trained healthcare staff collected rectal swabs and stools from participants. Rectal swabs were enriched in alkaline peptone water (APW) for 6-18 hours depending on patient admission time. Specimens were tested for *V. cholerae* by rapid diagnostic tests (RDTs) onsite, and by culture using standard methods (Appendix) at either an in-country reference laboratory, the Laboratoire Rodolphe Mérieux de l’Institut National de Recherche Biomédicale (INRB), in Goma (from October 2021 to September 2022), or at the onsite study laboratory (from September 2022 onward). As PCR for *V. cholerae* O1 was not available at either study laboratory in DRC, we shipped stool specimens (stool spotted on dry filter papers) for cases enrolled between October 14, 2021 and May 04, 2022 to Johns Hopkins University for PCR detection of toxigenic *V. cholerae* O1 following published methods.^25^

### Selection of cases and controls

Two cholera outbreaks occurred after mass vaccination, and we recruited cases during each outbreak, forming two distinct study periods (Figure 1). From 21 November 2022 to 24 January 2023, we retrospectively recruited controls for patients admitted to CTCs during the first outbreak (14 October 2021 to 10 March 2022), approximately 12-17 months after the second round of mass vaccination campaigns (Study Period 1). Between 17 October 2022 and 18 October 2023, we recruited controls for cases as they were admitted to the CTCs, 24-36 months after vaccination (Study Period 2).

**Figure 1.**
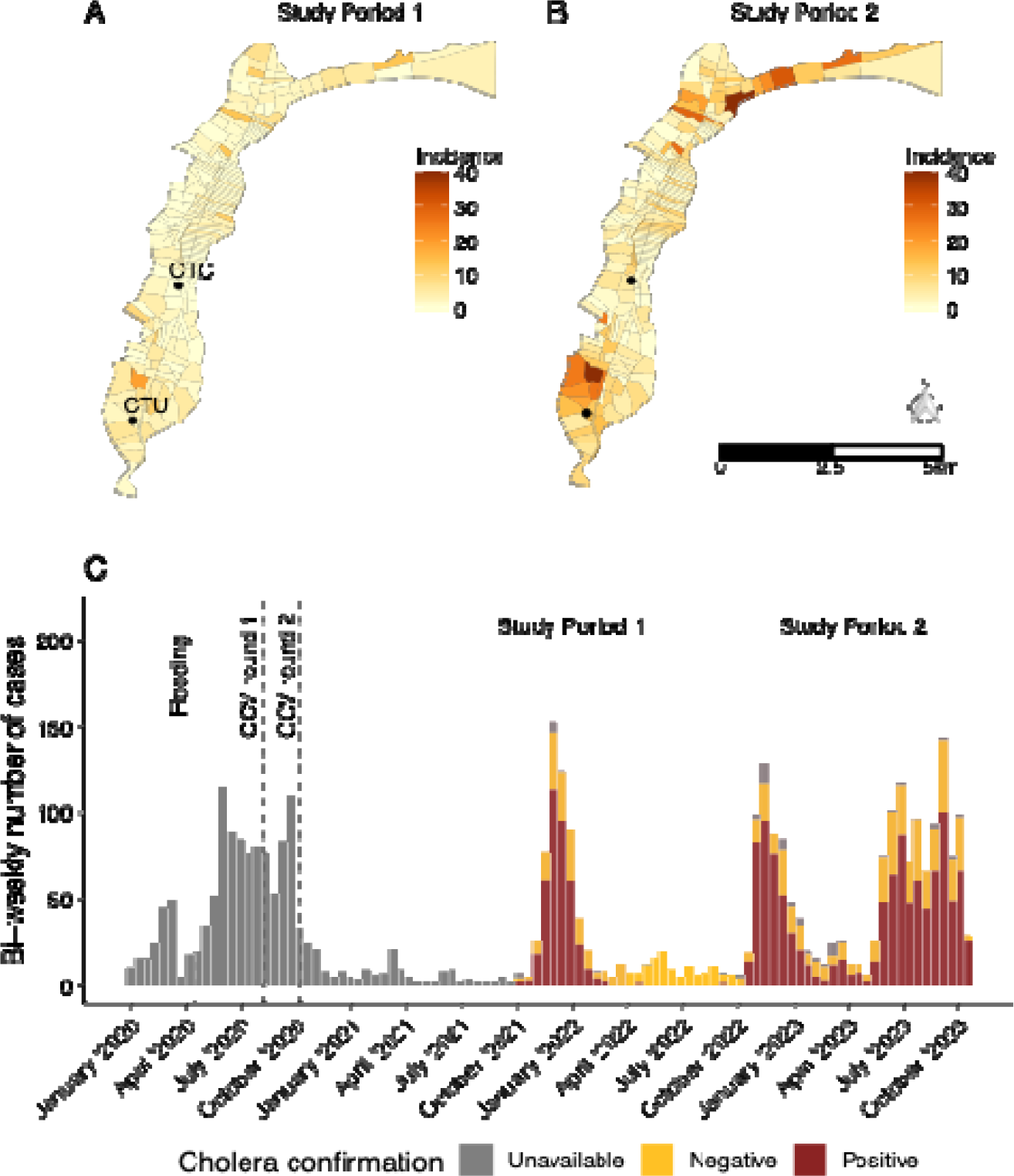
Number of cholera cases admitted to cholera treatment facilities in Uvira. Panels A (Study Period 1) and B (Study Period 2) illustrate the cumulative number of confirmed cases in each study period by neighborhood (avenue) across the city, with the locations of the two health facilities where patients were recruited shown as dots. There were 14 cases living in neighboring communities outside the city boundaries that were included in Study Period 2. The second outbreak (Study Period 2) started in the northern part of the city and spread to a refugee camp where many residents were admitted to the CTC but not included in the study as they were not living in Uvira at the time of vaccination. Panel C illustrates the epidemic curve of suspected and confirmed cholera cases admitted to the cholera treatment center (CTC) at the Uvira General Referral Hospital and the cholera treatment unit (CTU) at the Kalundu CEPAC health center. Cholera was confirmed by culture or PCR (Study Period 1), or by APW-enriched RDT and culture (Study Period 2). Among the 183 suspected cases that were detected before SP1 (in grey), 146 (79·8) were tested for *Vibrio cholerae* O1 by enriched RDT with 37 (25·3%) testing positive.

**Figure 2.**
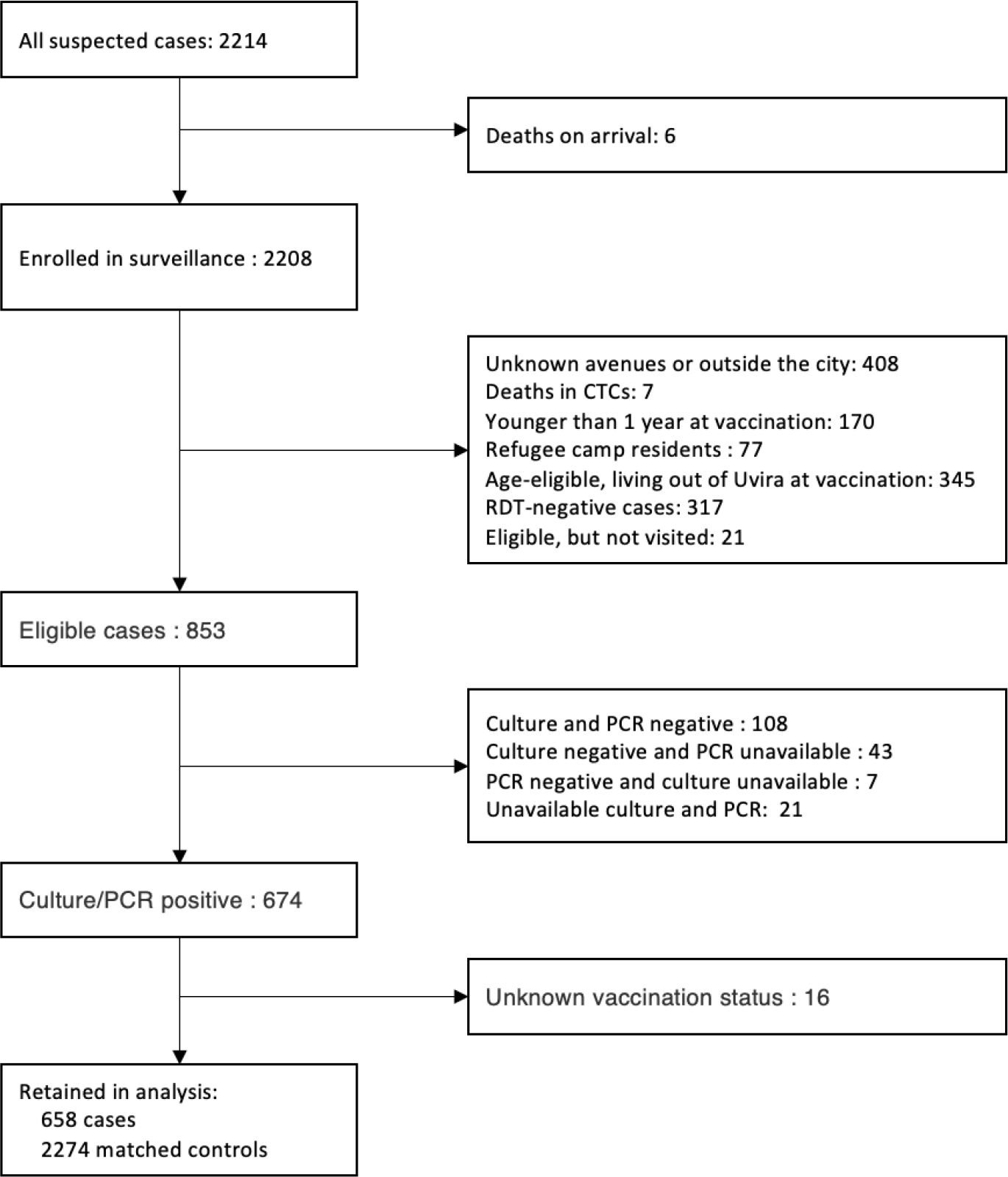
Flow chart of participant recruitment. Cases with unavailable culture results are those for which 1) suspected colonies were isolated, 2) with positive oxidase test at the field laboratory, 3) missing agglutination results due to an antiserum stockout, 4) and/or in which attempts to revitalize *Vibrio cholerae* O1 strains at the reference laboratory in Goma were unsuccessful.

Study Period 1 (SP1) included all consenting suspected cases who were at least 12 months old during the vaccination campaigns, living in Uvira for the two weeks prior to admission to the CTC and during the 2020 vaccination campaigns, and who tested positive for cholera by culture and/or PCR. We attempted to recruit four controls per case, using high-resolution satellite imagery (Appendix) to identify potential control households on the same avenue (smallest administrative unit in Uvira) as the case household. Control households were then selected by simple random spatial sampling of digitized residential structures. Controls were eligible for enrollment if: (1) they matched the case age group (1-4, 5-9, 10-19, 20-39, 40-59 or ≥60 years old) and sex, (2) had not been admitted for acute watery diarrhoea or cholera in the three years prior to the case admission, (3) were living in Uvira in the two weeks prior to case admission, (4) were living in Uvira at the time of the 2020 kOCV campaign and were eligible to be vaccinated, and (5) none of their household members reported being admitted to a formal health facility (as opposed to pharmacies, prayer homes or traditional healers) for acute watery diarrhoea or cholera in the four weeks prior to case admission (Table S1).

During Study Period 2 (SP2), the case definition included the same age and residence criteria as in SP1, but cases had to test positive with both APW-enriched rapid diagnostic test (RDT) and culture (performed at the onsite laboratory). We used enriched RDT results to help prioritize control recruitment due to limited human resources during the outbreak. In contrast to SP1, we conducted a home visit within three days of hospital discharge to investigate the living and WASH conditions in each case’s household and ascertain the vaccination status outside the hospital environment (as done with the controls). We excluded patients who died during hospitalization and those whose residence could not be found during home visits. As in SP1, four neighborhood controls were recruited from four randomly selected households near the case household, though during this study period we selected households using the ‘right-hand’ rule (Appendix). In addition to the recruitment criteria used in SP1, controls were eligible for enrolment in SP2 if their household matched that of the case by size (≤5 individuals, 6-10 individuals, and >10 individuals) and had at least one child below five years of age when the case household had one.

### Vaccination status ascertainment and potential confounding variables

Study staff administered structured questionnaires to all cases and controls (or their parent/guardian) to capture demographics, household conditions, potential confounding variables and vaccination status. Before asking each case or control whether they were vaccinated, study staff showed them photos of the vaccine vials and of someone taking the vaccine, in addition to explaining when and how the vaccines were delivered in Uvira, and how these may differ from other campaigns and routine vaccines. Participants reporting vaccination were asked the number of doses and when and where each one was taken. We also used vaccination cards to verify the vaccination status whenever possible. In SP1, vaccine-related questions were asked to cases in the clinic and in SP2 they were asked both in the clinic and a subsequent home visit. Any differences in the vaccination status reporting between the clinic and household interviews were solved through a third interview at the case’s household followed by a review of the data, discussion, and consensus within the study team. We identified potential confounders based on a causal directed acyclic graph developed before the start of the study and attempted to measure these through the interviews.

### Statistical analyses

The characteristics of case and control participants were compared using the standardized mean difference (SMD), which is the absolute difference in mean values of a variable between cases and controls divided by the pooled standard deviation. In addition, we calculated p-values from univariate conditional logistic regression models with case-control status as the dependent variable. In the primary analysis, we compared the odds of being vaccinated with a single kOCV dose between cases and controls using conditional logistic regression models. Those reporting to have received 2 or more doses were dropped from the primary analyses. The vaccine effectiveness (VE) was calculated as one minus the estimated odds ratio of having received a single dose of vaccine, between cases and controls. To produce consistent age-group specific and overall estimates of effectiveness across all ages, the conditional logistic regression model included an interaction term for age group (1-4 vs. ≥5) and the vaccination status. We derived the overall VE estimate based on a linear combination of parameters from the two age groups, weighted by the proportion of cases within each, and estimated simultaneous confidence intervals with the *ghlt* function in the *multcomp* R package.^26^ For continuous variables, we explored models using polynomials and restricted cubic splines, and compared them using Akaike Information Criterion (AIC). For combined estimates (SP1/2) and those from SP1, we incorporated age as a continuous variable, separately by the two age groups (with a quadratic term for the age group ≥ 5 years), to adjust for potential confounding. For SP2, estimates were adjusted for a set of potential confounders including a quadratic term for age, a cubic term for household size, household wealth index (as a continuous variable) derived from a principal component analysis of household assets ownership (Appendix), type of sanitation facility, whether the participant used a toilet shared by multiple households compared to using a private toilet, drinking water sources, and availability of a hand washing facility and soap. We also fit three alternative models with different sets of covariates to assess the robustness of the estimates (Appendix). In a secondary analysis, we estimated the VE for at least one dose and two doses of kOCV compared to the unvaccinated group. While we primarily relied on point estimates and 95% confidence intervals (CI) to assess the weight of evidence, we considered p-values statistically significant when they were less than 0.05. Analyses were performed in R (version 4.3.1).

### Ethical considerations

Ethical approvals were obtained from Institutional Review Boards of the Johns Hopkins Bloomberg School of Public Health (IRB00015785), the London School of Hygiene & Tropical Medicine (25365) and the École de Santé Publique at the University of Kinshasa (ESP/CE/65/2021). Written informed consent was obtained from all participants ≥18 years, with written assent from those <18 years in addition to written consent from their parent/guardian.

### Role of the funding source

The funders of the study had no role in study design, data analysis, interpretation, preparation of this manuscript, or the decision to publish.

## Results

### Cholera incidence in Uvira and recruitment of study participants

We recruited 658 unique confirmed cholera cases and 2,274 matched controls during the two study periods (Figure 1), with 61·1% of cases enrolled in the second period. The median age of participants at the time of the vaccination campaigns was 14·9 (interquartile range 6·3, 33·8) years and 15·1% were under five years old (Table 1). Almost sixty-two percent of cases were recorded as severely dehydrated on admission. Cases were significantly older during the first study period compared to the second period (p=0·005), though they all had similar dehydration status (Table S2). Of the 537 culture positive isolates, 390 (72·6%) were serotype Ogawa, the rest were Inaba, including 19·3% (n=26/135) Inaba in SP1 and 30·1% (n = 121/402) Inaba in SP2.

**Table 1.** Characteristics of participants by case and control status.

| Characteristic | Overall,<br>N = 2,932 | Cases,<br>N = 658 | Controls,<br>N = 2,274 | P value | SMD |
| --- | --- | --- | --- | --- | --- |
| Age at vaccination,<br>median (IQR) | 14.9 (6.3,<br>33.8) | 14.1 (6.0, 33.8) | 15.1 (6.4,<br>33.7) | 0.0019 | 0.005 |
| Age group at<br>vaccination (years) |  |  |  |  | 0.122 |
| 1-4 | 443 (15.1%) | 99 (15.0%) | 344 (15.1%) | Reference |  |
| 5-9 | 552 (18.8%) | 146 (22.2%) | 406 (17.9%) | 0.0080 |  |
| 10-19 | 764 (26.1%) | 159 (24.2%) | 605 (26.6%) | 0.7183 |  |
| 20-39 | 606 (20.7%) | 124 (18.8%) | 482 (21.2%) | 0.8436 |  |
| 40-59 | 401 (13.7%) | 91 (13.8%) | 310 (13.6%) | 0.4146 |  |
| ≥60 | 166 (5.7%) | 39 (5.9%) | 127 (5.6%) | 0.4495 |  |
| Sex |  |  |  |  | 0.001 |
| Female | 1,503<br>(51.3%) | 337 (51.2%) | 1,166 (51.3%) | Reference |  |
| Male | 1,429<br>(48.7%) | 321 (48.8%) | 1,108 (48.7%) | 0.8577 |  |
| Vaccination status |  |  |  |  | 0.277 |
| Not Vaccinated | 1,732<br>(59.1%) | 452 (68.7%) | 1,280 (56.3%) | Reference |  |
| One Dose | 851 (29.0%) | 133 (20.2%) | 718 (31.6%) | <0.0001 |  |
| Two Doses | 349 (11.9%) | 73 (11.1%) | 276 (12.1%) | 0.0110 |  |
| Vaccination card<br>available | 163 (13.6%) | 29 (14.1%) | 134 (13.5%) | 0.1092 | 0.017 |

P values are obtained from univariable conditional logistic regression models.

Overall, 20·2% of cases reported receiving one dose of kOCV and 68·7% were unvaccinated. In comparison, 31·6% of matched controls reported receiving a single dose of kOCV and 56·3% were unvaccinated. Only 13·6% of vaccinated participants were able to show a vaccination card and 11·9% reported having received two doses of the vaccine (Table 1).

Additional data on socio-demographic and household characteristics were collected in SP2 (Table 2, Table S4). Cases were more likely to use toilets shared by multiple households than controls (OR 1·41; 95% CI 1·11-1·79) and were less likely to live in houses with electricity (OR 0·72; 95% CI 0·56–0·92). Cases were also more likely to live in households with lower wealth index, indicating higher level of poverty (OR 0·58; 95% CI 0·44–0·76) than controls. Although likely an artifact of hygiene kit distribution that focused on case households, we found that cases were more likely to live in households with soap and water available for handwashing (OR 1·86; 95% CI 1·40–2·49).

**Table 2.**
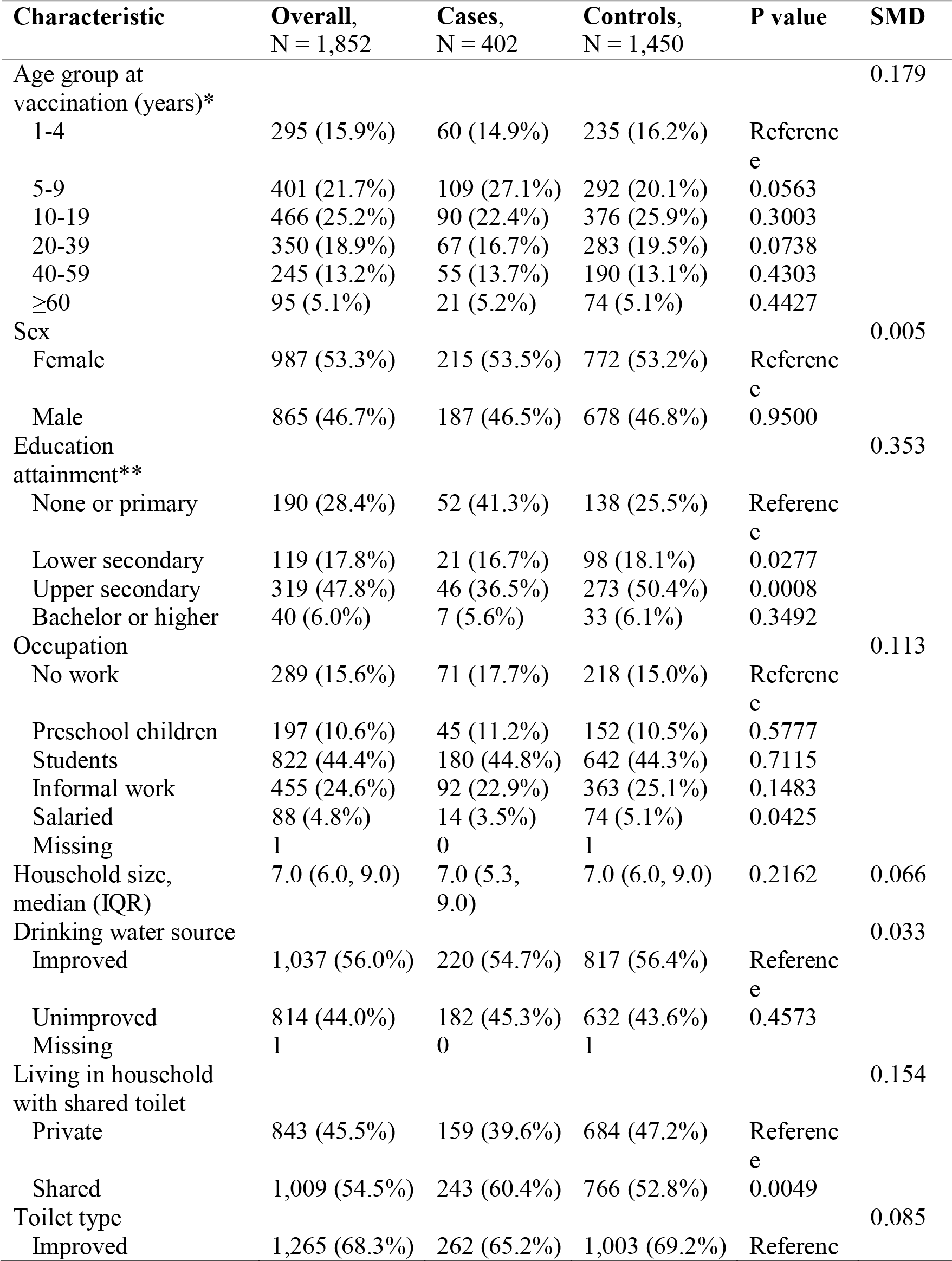
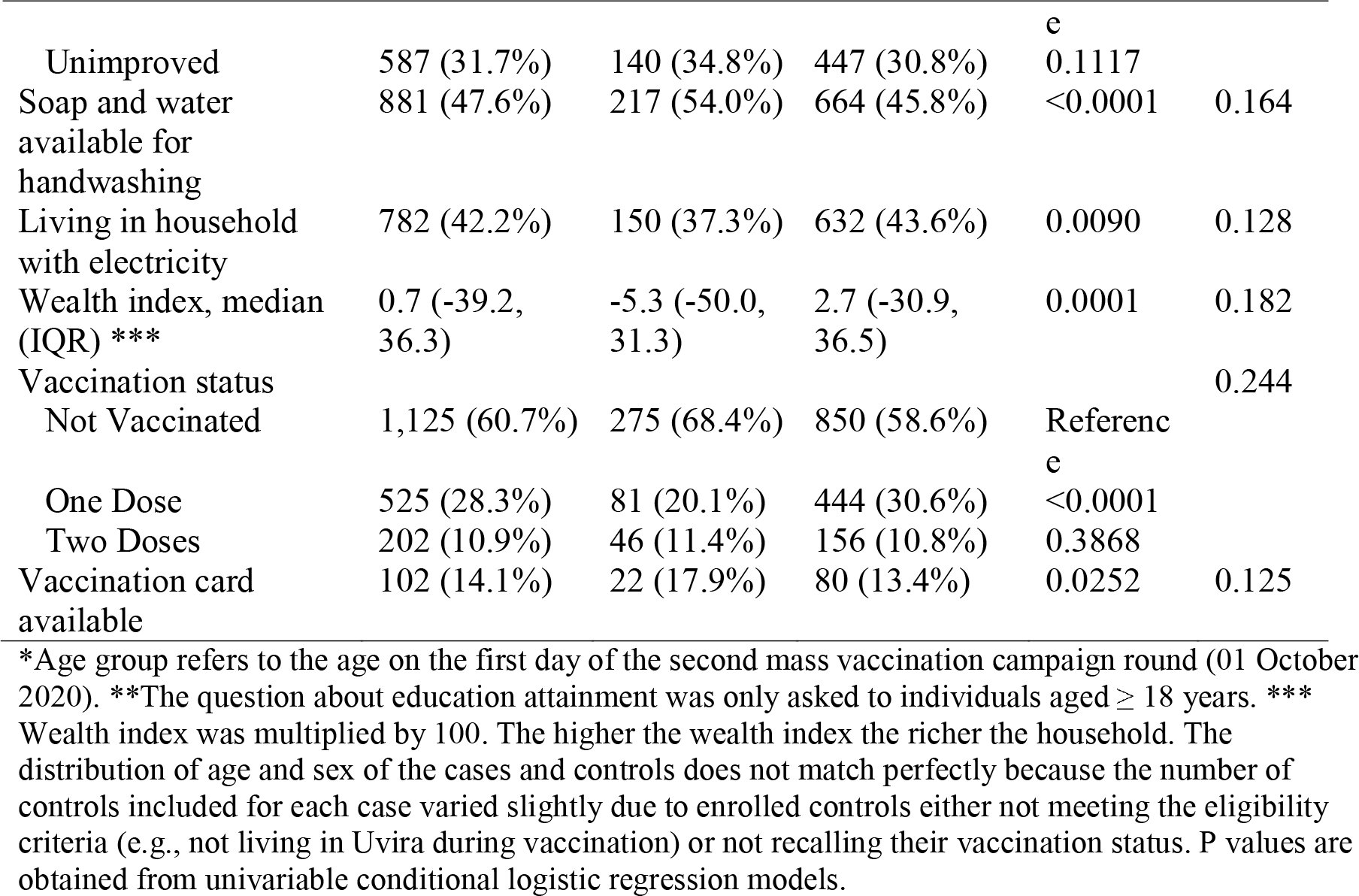
Characteristics of study participants in the vaccine effectiveness analysis 24-36 months after vaccination (Study Period 2)

Combining data from both study periods, 12-36 months post-vaccination, we estimated an unadjusted single-dose VE of 47·8% (95% Confidence Interval [CI] 34.6–58·4) and after adjustment for potential confounders, the VE was 48·2% (95% CI 34·8–58·8).

In SP1, 12–17 months after vaccination, we estimated an unadjusted and adjusted single-dose VE of 54·4% (95% CI: 34·4–68·3) and 52·7% (95% CI: 31·4–67·4). In SP2, 24–36 months after vaccination, we estimated an unadjusted VE of 43·2% (95% CI: 24·0–57·6) and an adjusted VE of 45·5% (95% CI: 25·8–60·0). The adjusted VE for 1–4-year-olds was 73·5% (95% CI: 28·9– 90·1) in SP1 sharply declining to 33·1% (95% CI: -30·0–65·6) in SP2 and the confidence intervals span the null (Table 3).

**Table 3.** Effectiveness of a single dose of oral cholera vaccine, 12-17 months and 24-36 months after vaccination campaigns.

| Population | Cases (effective n*) | Controls (effective n*) | Unadjusted VE (95% CI) | Adjusted VE** (95% CI) |
| --- | --- | --- | --- | --- |
| **12-36 months after vaccination** |  |  |  |  |
| Overall | 573 (419) | 1998 (763) | 47.8% (34.6–58.4) | 48.2% (34.8–58.8) |
| 1-4 years | 96 (61) | 263 (129) | 52.4% (22.5–70.8) | 49.8% (15.6–70.2) |
| $\geq 5$ years | 463 (332) | 1534 (584) | 46.7% (31.8–58.4) | 47.8% (32.8–59.5) |
| **12-17 months after vaccination (Study Period 1)** |  |  |  |  |
| Overall | 219 (170) | 704 (300) | 54.4% (34.4–68.3) | 52.7% (31.4–67.4) |
| 1–4 years | 34 (23) | 83 (44) | 68.3% (19.6–87.5) | 73.5% (28.9–90.1) |
| ≥5 years | 179 (141) | 568 (243) | 50.9% (28.0–66.6) | 46.9% (21.0–64.3) |
| **24–36 months after vaccination (Study Period 2)** |  |  |  |  |
| Overall | 354 (249) | 1294 (463) | 43.2% (24.0–57.6) | 45.5% (25.8–60.0) |
| 1–4 years | 62 (38) | 180 (85) | 42.8% (–2.2–68.0) | 33.1% (–30.0–65.6) |
| ≥5 years | 284 (191) | 966 (341) | 43.4% (21.7–59.0) | 48.3% (27.2–63.3) |

\*Effective number of cases and controls is the sample size that effectively contributes to estimates of effectiveness in the conditional logistic regression models. Those participants in matched case-control sets where all people have the same vaccination status do not contribute to the estimates, nor do those who either do not know their vaccination status or the number of doses they received. \*\*In SP1 and in analyses combining data from both study periods, we only adjusted for age as a continuous variable

In secondary analyses, we estimated the adjusted cumulative VE across both study periods for at least one dose of kOCV to be 45·4% % (95% CI: 33·2–55·3, Table S8). We were unable to reliably estimate two-dose VE due to a limited effective sample size, with post-hoc power calculations suggesting we had 50·9% power to detect significant vaccine protection even when combining data from both study periods and 5.1% power in young children (Table S9, Table S10).

## Discussion

We found that a single dose of Euvichol-Plus^®^ kOCV provided protection against cholera for at least the 36-month period post-vaccination. Our study provides unique, policy-relevant insights into kOCV protection as we estimated single-dose effectiveness from the only available and most widely used cholera vaccine today, including estimates of effectiveness for 1–4-year-olds and at discrete time windows after vaccination. Our results suggest that at least in cholera endemic areas like Uvira, the use of one kOCV dose may provide significant protection on the scale of years rather than just months.

To date, one randomized trial^17,18^ and seven observational studies have included estimates of one-dose protection of kOCVs.^7–9,13–15,27^ Most of these studies have been short-term estimates of protection measured for just a few months after vaccination and showed similar levels of protection to two doses on this timescale. Two notable exceptions where protection was measured over a longer period include a randomized trial in Bangladesh and a case-control study in Haiti, both using Shanchol^®^. The Bangladesh trial was conducted for two years and estimated 54% (95% CI: 16 to 75) efficacy in the first year and 67% (43 to 81) in the second year post-vaccination among those five years and older.^17^ Secondary analyses from a case-control study in Haiti predicted that a single dose of kOCV (Shanchol^®^) would confer 58% (95% CI: 4 to 82%) protection at 16 months post-vaccination, with the confidence intervals including zero protection from 17-months post-vaccination and onward.^14^ Our results are consistent with these previous studies in showing significant protection for the overall population for longer than a year, though it is important to note that Uvira, like Haiti (at the time of the study) and Bangladesh, is endemic for cholera so the first dose may have acted as a booster for previously exposed individuals. Furthermore, it is possible that previous exposures to enterotoxigenic *Escherichia coli*, which has been identified among diarrhoea cases in Uvira and has a antigenically similar toxin to toxigenic *V. cholerae*, could contribute to background immunity to severe disease in these settings.^28^ More work is needed to characterize the epidemiologic settings where one dose may provide comparable levels of protection to the full regimen, and might include leveraging historic incidence rates of cholera or, perhaps, population-level immunologic measures of previous exposures.^29^

Before this study, only one estimate of single-dose protection of a kOCV in 1–4-year-olds had been published. This trial, in Bangladesh, suggested that young children did not benefit from a single dose of Shanchol^®^, even during the first six months post-vaccination.^17,18^ In contrast, we found evidence that the population under five years old in Uvira benefited from similar levels of protection to those five and above at 12-17 months post-vaccination, however the point estimates dropped substantially in the third year after vaccination with confidence intervals spanning the null. This observation is in line with studies showing similar levels of protection after natural infection between the two age groups.^30^ Conflicting estimates between young children and older individuals have also been observed in kOCV studies with the full dose regimen, though there are only a handful of studies that present age-stratified estimates. Although most studies have found lower effectiveness in young children, the difference in protection has been highly variable with large uncertainty (e.g., ranging from no apparent difference in Vietnam after 10 months,^31^ to 73% lower protective efficacy among children in Bangladesh over 6 months post-vaccination^18^). The contrast of our estimates with other studies could be explained by several factors including pre-existing population immunity and season of vaccination as shown with other diseases,^32^ differences both in gut microbiota composition and in prevalence of enteropathy, however, as in all observational studies we cannot rule out confounding and selection bias.^33,34^ Additional observational, and ideally randomized trials, would help shed further light on protection of one dose of kOCV among young children.

While the focus of the study was to understand the direct effectiveness of kOCV, our analysis of potential risk factors highlighted key differences between cases and matched controls. Our results confirm the associations between cholera and markers of poverty, like using a shared latrine, using an unimproved source for drinking water and not having electricity in the household. While kOCV provides protection against cholera, tackling fundamental risk factors like access to safe water and sanitation are needed to sustainably control the disease.

This study comes with several limitations. First, like many previous kOCV effectiveness studies, the vaccination status was self-reported,^7,14,15^ and only 29 cases (of 206 reporting to be vaccinated) were able to provide a vaccination card. Measurements of the number of doses received months to years after a mass vaccination campaign is prone to recall bias, particularly in a place like Uvira where mass vaccination campaigns with different antigens are common. To minimize biases in classification of vaccination status, we used visual aids and a series of structured questions, and hospital and study staff reassured patients that their responses to the study questions would in no way affect their care. Though enumerators were not blinded to the vaccination status of the cases while enrolling their matched controls, the vaccine coverage among controls was similar to that measured in community coverage surveys (Table S12). Furthermore, sensitivity analyses restricted to only those with a vaccination card revealed similar effectiveness estimates (Table S5). There were slight differences in the protocols of the case-control study in each study period, challenging the interpretability of the joint estimates from both periods. However, in sensitivity analyses simulating similar diagnostic criteria for cases in SP1 and SP2, our qualitative findings remained consistent (Table S11). The retrospective recruitment of controls for cases admitted during the SP1 precluded adjustment for individual and household factors that may have influenced cholera disease risk or vaccine acceptability.

However, such factors are unlikely to have significantly influenced the magnitude of our VE estimates as we observed in the SP 2 (Table S6 and S7) and other cholera VE studies conducted elsewhere.^7,14,15^ The retrospective nature of recruitment of controls in SP1 could also have led to differential recall of vaccination status among cases and their controls. Even in SP2, our VE estimates could still be confounded by unmeasured factors. Although the number of cases under five years of age was higher than in most published VE studies, our sample size in this important age group was still small and led to wide confidence intervals around VE estimates (Table S2 and S10). Finally, we were unable to obtain reliable estimates of two dose protection partly because few cases reported receipt of two doses of kOCV due to low vaccination coverage in the population, and potentially because of uncertainty in the reporting of more than one dose of kOCV (Tables S2, S3 S9 and Figure S2).

Our findings, combined with data from dose-interval studies conducted in Cameroon and Zambia^35,36^, suggest that providing a second dose a year or more after the first could lead to better and longer lasting protection against cholera than the current two-dose series, at least among older children and adults. While more data are needed across different settings and for longer periods of time, our study extends the current evidence base on protection from a single-dose of kOCV, and more specifically on protection from Euvichol-Plus^®^, the most widely used cholera vaccine available today.

## Data Availability

All data produced in the present study are available upon reasonable request to the authors

## Declarations

### Funding

This work was supported by the Wellcome Trust and Gavi (GAVI-RFP-2019-062).

### Authors’ contributions

Conceptualization: EBM, ASA, JK. Data curation: EBM, JDH, HX, CH, PMB, ASA. Formal analysis: EBM, ASA. Data interpretation: EBM, ASA, JK, JL, EL. Funding acquisition: ASA, KG, OC. Investigation: EBM, PMB, CH, HX, JDH, BBR, MI, ETT, FK, TBM, AD, EL, DL, JK, ASA. Methodology: EBM, ASA, CH, JK. Project Administration: EBM, PMB, MD, DMB, HKM, CH, ASA, KG. Resources: PWO, JMS, DMB, AD, JK, OL. Software: EBM, JDH, HX, ASA. Supervision: ASA, JK. Validation: EBM, ASA. Visualization - EBM, ASA. Writing - original draft - EBM, ASA. Writing: review & editing: all authors.

## Acknowledgements

We are thankful to Faraja Masema Lulela, Joël Faraja Zigashane Mashauri, Jean-Marie Masugamuhanya Cirhonda, the CTCs nurses and field investigators for their support for the data collection. We also thank the Head of the Uvira Health Zone, Dr Panzu Nimi, and the Director of the Uvira General Referral Hospital, Dr Salomon Mashupe, for their operational and administrative support throughout this study. We would also like to thank the MSF GIS Centre, including Frederic Ham and Edith Rogenhofer, for their assistance with acquiring satellite imagery data and Ahmad Alobaidi for assistance with dwelling extractions from this imagery.

## Data sharing

Code and data from this study are available at https://github.com/HopkinsIDD/uvira_onedose_ocv_ve.

## Notes

### Competing Interest Statement

The authors have declared no competing interest.

### Author Declarations

Ethical approvals were obtained from Institutional Review Boards of the Johns Hopkins Bloomberg School of Public Health (IRB00015785), the London School of Hygiene & Tropical Medicine (25365) and the Ecole de Sante Publique at the University of Kinshasa (ESP/CE/65/2021).

### Summary of Updates

Updated version and new data after peer rivew.

## References

1 Deshpande A, Miller-Petrie MK, Lindstedt PA, et al. Mapping geographical inequalities in access to drinking water and sanitation facilities in low-income and middle-income countries, 2000–17. The Lancet Global Health 2020; 8: e1162–85.

2 World Health Organization. Global Task Force on Cholera Control. Ending Cholera, a Global Roadmap to 2030. World Health Organization, 2017.

3 Bi Q, Ferreras E, Pezzoli L, et al. Protection against cholera from killed whole-cell oral cholera vaccines: a systematic review and meta-analysis. Lancet Infect Dis 2017; 17: 1080–8.

4 Ali M, Qadri F, Kim DR, et al. Effectiveness of a killed whole-cell oral cholera vaccine in Bangladesh: further follow-up of a cluster-randomised trial. Lancet Infect Dis 2021; 21: 1407–14.

5 Burki T. Addressing the shortage of cholera vaccines. Lancet Infect Dis 2022; 22: 1674–5.

6 Odevall L, Hong D, Digilio L, et al. The Euvichol story - Development and licensure of a safe, effective and affordable oral cholera vaccine through global public private partnerships. Vaccine 2018; 36: 6606–14.

7 Luquero FJ, Grout L, Ciglenecki I, et al. Use of Vibrio cholerae vaccine in an outbreak in Guinea. N Engl J Med 2014; 370: 2111–20.

8 Azman AS, Parker LA, Rumunu J, et al. Effectiveness of one dose of oral cholera vaccine in response to an outbreak: a case-cohort study. Lancet Glob Health 2016; 4: e856–63.

9 Grandesso F, Kasambara W, Page A-L, et al. Effectiveness of oral cholera vaccine in preventing cholera among fishermen in Lake Chilwa, Malawi: A case-control study. Vaccine 2019; 37: 3668–76.

10 Sialubanje C, Kapina M, Chewe O, et al. Effectiveness of two doses of Euvichol-plus oral cholera vaccine in response to the 2017/2018 outbreak: a matched case–control study in Lusaka, Zambia. BMJ Open 2022; 12: e066945.

11 World Health Organization. Weekly epidemiological record. World Health Organization, 22 SEPTEMBER 2023, 98th YEAR https://iris.who.int/handle/10665/372986 (accessed Oct 17, 2023).

12 World Health Organization. Shortage of cholera vaccines leads to temporary suspension of two-dose strategy, as cases rise worldwide. 2022; published online Oct 19. https://www.who.int/news/item/19-10-2022-shortage-of-cholera-vaccines-leads-to-temporary-suspension-of-two-dose-strategy--as-cases-rise-worldwide (accessed March 24, 2023).

13 Khatib AM, Ali M, von Seidlein L, et al. Effectiveness of an oral cholera vaccine in Zanzibar: findings from a mass vaccination campaign and observational cohort study. Lancet Infect Dis 2012; 12: 837–44.

14 Franke MF, Ternier R, Jerome JG, Matias WR, Harris JB, Ivers LC. Long-term effectiveness of one and two doses of a killed, bivalent, whole-cell oral cholera vaccine in Haiti: an extended case-control study. Lancet Glob Health 2018; 6: e1028–35.

15 Ferreras E, Blake A, Chewe O, et al. Alternative observational designs to estimate the effectiveness of one dose of oral cholera vaccine in Lusaka, Zambia. Epidemiol Infect 2020; 148: e78.

16 Ivers LC, Hilaire IJ, Teng JE, et al. Effectiveness of reactive oral cholera vaccination in rural Haiti: a case-control study and bias-indicator analysis. Lancet Glob Health 2015; 3: e162–8.

17 Qadri F, Ali M, Lynch J, et al. Efficacy of a single-dose regimen of inactivated whole-cell oral cholera vaccine: results from 2 years of follow-up of a randomised trial. Lancet Infect Dis 2018; 18: 666–74.

18 Qadri F, Wierzba TF, Ali M, et al. Efficacy of a Single-Dose, Inactivated Oral Cholera Vaccine in Bangladesh. N Engl J Med 2016; 374: 1723–32.

19 Bugeme PM, Xu H, Hutchins C, et al. Cholera deaths during outbreaks in Uvira, Eastern Democratic Republic of Congo, September 2021-January 2023. bioRxiv. 2023; published online June 3. DOI:10.1101/2023.05.26.23290528.

20 Schyns C, Fossa A, Mutombo-Nfenda, et al. Cholera in Eastern Zaire, 1978. Ann Soc Belg Med Trop 1979; 59: 391–400.

21 Jeandron A. Tap water access and its relationship with cholera and other diarrhoeal diseases in an urban, cholera-endemic setting in the Democratic Republic of the Congo. 2020; published online Dec 12. DOI:10.17037/PUBS.04659288.

22 Jeandron A, Cumming O, Kapepula L, Cousens S. Predicting quality and quantity of water used by urban households based on tap water service. npj Clean Water 2019; 2: 1–9.

23 Gaiffe M, Dross C, Malembaka EB, Ross I, Cumming O, Gallandat K. A fuzzy inference-based index for piped water supply service quality in a complex, low-income urban setting. Water Res 2023; : 120316.

24 Gallandat K, Macdougall A, Jeandron A, et al. 1 Improved water supply infrastructure to reduce acute diarrhoeal diseases and 2 cholera in Uvira, Democratic Republic of the Congo: results and lessons learned 3 from a pragmatic trial. https://osf.io/na47d/download (accessed July 23, 2023).

25 Debes AK, Ateudjieu J, Guenou E, et al. Clinical and Environmental Surveillance for Vibrio cholerae in Resource Constrained Areas: Application During a 1-Year Surveillance in the Far North Region of Cameroon. Am J Trop Med Hyg 2016; 94: 537–43.

26 Hothorn T, Bretz F, Westfall P. Simultaneous inference in general parametric models. Biom J 2008; 50: 346–63.

27 Wierzba TF, Kar SK, Mogasale VV, et al. Effectiveness of an oral cholera vaccine campaign to prevent clinically-significant cholera in Odisha State, India. Vaccine 2015; 33: 2463–9.

28 Clemens JD, Sack DA, Harris JR, et al. Cross-Protection by B Subunit-Whole Cell Cholera Vaccine Against Diarrhea Associated with Heat-Labile Toxin-Producing Enterotoxigenic Escherichia coli: Results of a Large-Scale Field Trial. J Infect Dis 1988; 158: 372–7.

29 Azman AS, Lessler J, Luquero FJ, et al. Estimating cholera incidence with cross-sectional serology. Sci Transl Med 2019; 11. DOI:10.1126/scitranslmed.aau6242.

30 Ali M, Emch M, Park JK, Yunus M, Clemens J. Natural Cholera Infection–Derived Immunity in an Endemic Setting. J Infect Dis 2011; 204: 912–8.

31 Trach DD, Clemens JD, Ke NT, et al. Field trial of a locally produced, killed, oral cholera vaccine in Vietnam. Lancet 1997; 349: 231–5.

32 Moore SE, Collinson AC, Fulford AJC, et al. Effect of month of vaccine administration on antibody responses in The Gambia and Pakistan. Trop Med Int Health 2006; 11: 1529–41.

33 Zimmermann P, Curtis N. Factors That Influence the Immune Response to Vaccination. Clin Microbiol Rev 2019; 32. DOI:10.1128/CMR.00084-18.

34 Porras AM, Shi Q, Zhou H, et al. Geographic differences in gut microbiota composition impact susceptibility to enteric infection. Cell Rep 2021; 36: 109457.

35 Mwaba J, Chisenga CC, Xiao S, et al. Serum vibriocidal responses when second doses of oral cholera vaccine are delayed 6 months in Zambia. Vaccine 2021; 39: 4516–23.

36 Ateudjieu J, Sack DA, Nafack SS, et al. An Age-stratified, Randomized Immunogenicity Trial of Killed Oral Cholera Vaccine with Delayed Second Dose in Cameroon. Am J Trop Med Hyg 2022; 107: 974–83.

