## Appendix for "Effectiveness of one dose of killed oral cholera vaccine in an endemic community in the Democratic Republic of the Congo: A matched case-control study"

**Web Appendix**

**Content**

### 1. Supplementary Methods

### 1.1. Cholera confirmation

For each patient, we used a new, untreated, plastic bucket to collect fresh stools to avoid the impact of residual chlorine used for cleaning in CTCs on culture sensitivity. Stool specimens were therefore collected using two methods: rectal swabs which were taken by healthcare staff and a stool specimen container was used to aliquot stool from the collection bucket. Rectal swab specimens were enriched in alkaline peptone water (APW) for 6–18 hours (depending on patient admission time), and both APW-enriched and fresh stool specimens were tested with Crystal VC Rapid Dipstick (O1/O139 or O1-only, Arkray Healthcare Pvt Ltd, Gujarat, India) tests at the bedside for cases admitted before March 27, 2022, and in the onsite lab for APW-enriched tests for cases admitted after this date.

The same culture protocol was observed in both the onsite and the reference laboratory. APW-enriched samples were plated directly onto thiosulfate citrate bile salt sucrose (TCBS) agar and incubated at 37°C for 18-24 hours. Suspected *V. cholerae* colonies were subcultured onto TCBS. After 18-24 hours of incubation, the suspected *V. cholerae* isolates were further subcultured on Tryptone Soya Agar (TSA) and incubated at 37°C for 18-24 hours. Colonies from the TSA agar were tested for oxidase activity using Oxidase strips (Merck Millipore, UK) and oxidase-positive isolates were further tested for autoagglutination with saline solution. Non-autoagglutinating isolates were tested for agglutination using Polyvalent-O1, Ogawa and Inaba antisera (MAST group Ltd., UK).

#### 1.1. Wealth index

The socio-economic position of households was characterized by conducting a principal component analysis on household assets and housing characteristics to create a wealth index. This index was based on ownership of transportation means (bicycle, motorcycle, three wheeler, car or pirogue), domestic animals (duck or chicken, goat, pig, cattle), mobile phone, computer, tablet, radio, television, satellite dish, refrigerator, and on housing structure (whether the house is paved with cement, tiles or slab, or not; weather the house walls are in tiles, bricks, cement/slab or not), and whether the house has a permanent source of electricity (that is, electricity from the national electricity grid or fixed solar panels, or not). We extracted three components and used the varimax rotation method. A wealth index was generated as the score of the first principal component that explains the largest proportion of the total variance. Similar approaches have been extensively used in Demographic and Health Surveys and contexts, especially where reliable data on income or household consumption expenditures are not available.^3^

#### 1.3. Random spatial selection of control households and selection of study participants

In both study periods, we attempted to randomly recruit four controls for each case. In Study Period 1 (12-17 months post-vaccination), control households were selected in the case’s *avenue* (the lowest administrative level in Uvira) of residence by simple random spatial sampling of potential residential buildings identified from a high-resolution satellite image acquired between February 26 and March 16, 2021 (Airbus, Pléiades 1B sensor). Through an iterative process of machine learning and manual review of imagery, 59,065 structures were identified as potential residential buildings within the Uvira city boundaries. We excluded 495 structures that had large areas (greater than 500 m^2^) as we assumed they were unlikely to be residential.

We used the *OsmAnd* mobile app to geo-locate and identify the sampled structures. A GPX file containing the structure IDs and GPS coordinates was uploaded to *OsmAnd* to track the selected structures. If the dwelling structure was multi-story, the household units were numbered from bottom to top and one household was randomly selected using the *Pretty Random - Random Number* mobile application. If the structure was a single-story building with multiple residential units, the latter were numbered starting with the unit closest to the GPS point, and one unit was chosen at random. When the sampled structure was not residential, field investigators were asked to approach and enroll the residential structure nearest to the GPS point within a radius of 20 meters. In the case there was no residential structure within 20 meters of the point, the “right hand rule” was used for the selection of control households. The investigator stood in front of the sampled structure, then selected a random number, X, between one and five, using *Pretty Random - Random Number*. They then walked to the X^th^ residential structure in the right-hand direction. In the event of a refusal or presence of a non-residential structure, this process was repeated until a consenting household was found. Once the dwelling structure was found, a householder or his representative was identified. If there was no head of household or adult representative at the first visit, investigators were asked to revisit the household up to two more times during the survey period.

In Study Period 2, controls were selected in the case’s neighborhood, using the right-hand rule described above, starting from the case’s household. That was possible in this study period because the exact address of the case’s household was known through home visits. We excluded patients coming from a camp of refugees who were living in two temporary and shared structures (one for women and boys below 10 years of age, and another for men aged at least 10 years), because 1) it was not possible to identify unique households within the camp, 2) residents of the camps were not yet living in Uvira at the time of vaccination, and 3) they do not share the same risk factors as the local community in Uvira. Inclusion and exclusion criteria applied in both study periods are summarized in Table S1.

**Table S1. Summary of inclusion and exclusion criteria**

| **Study Period 1** | Cases | Controls |
| --- | --- | --- |
| Household inclusion | Household in which the case has been living for at least two weeks before admission to the CTC. | 1. No household members had reported being admitted to a health facility for acute watery diarrhea in the 4 weeks before the date of the case's CTC admission. 2. Have at least one eligible participant |
| Individual inclusion | 1. Consenting suspected cases living in Uvira for at least two weeks prior to admission 2. Positive cholera culture and/or PCR 3. Aged at least one year and living in Uvira at the time of vaccination. 4. Residential address in one of the avenues in the city of Uvira | 1. Same age group (1-4, 4-9, 10-19, 20-39, 40-59, ≥60 years) and same sex as the case  2. Have been living in that household for the two weeks preceding admission of the matched case to the CTC  3. Have not been admitted to a health facility for acute watery diarrhea or cholera in the past 3 years prior to the case’s CTC admission,  4. Aged at least one year and have been living in Uvira at the time of vaccination.  5. If multiple individuals were eligible to be matched controls, study staff selected one at random to attempt to enroll into the study |
| Individual exclusion | 1. Unknown residential address or residential address outside the city boundaries 2. Unknown vaccination status | Unknown vaccination status |
| **Study Period 2** |  |  |
|  | Household located in Uvira in which the case has been living for at least two weeks before her/his admission to CTC | 1. Similar household size as the case (≤5 individuals, 6-10 individuals, and >10 individuals). 2. Have at least one child below five years of age when the case household had one child of this age. 3. No household members had reported being admitted to a health facility for acute watery diarrhea in the 4 weeks before the date of the case's CTC admission. 4. Have at least one eligible participant |
| Individual Inclusion | 1. Consent of suspected cases testing positive by both APW-enriched RDT and culture (onsite). 2. Living in Uvira for at least two weeks prior to admission 3. Aged at least one year and living in Uvira at the time of vaccination. 4. Residential address in one of the avenues in the city of Uvira | 1. Living in the sampled household control for at least two weeks 2. Not having been admitted to health facility for acute watery diarrhea or cholera in the past three year and 3. Same age group (1-4, 4-9, 10-19, 20-39, 40-59, ≥60 years) and sex (for participants aged ≥ 5 years) as the corresponding case, 4. Aged at least one year and have been living in Uvira at the time of vaccination. 5. In the presence of multiple eligible household members, one was selected randomly with a random number generator. |
| Individual exclusion | 1. Resident of a camp of refugees who were living in shared temporary structures (community in Uvira. 2. Patients who died during hospitalization 3. Patients in transit in Uvira, 4. Patients whose residence could not be found during home visits. 5. Unknown vaccination status | Unknown vaccination status |

### 2. Supplementary Results

#### 2..1. Participants by Study Period

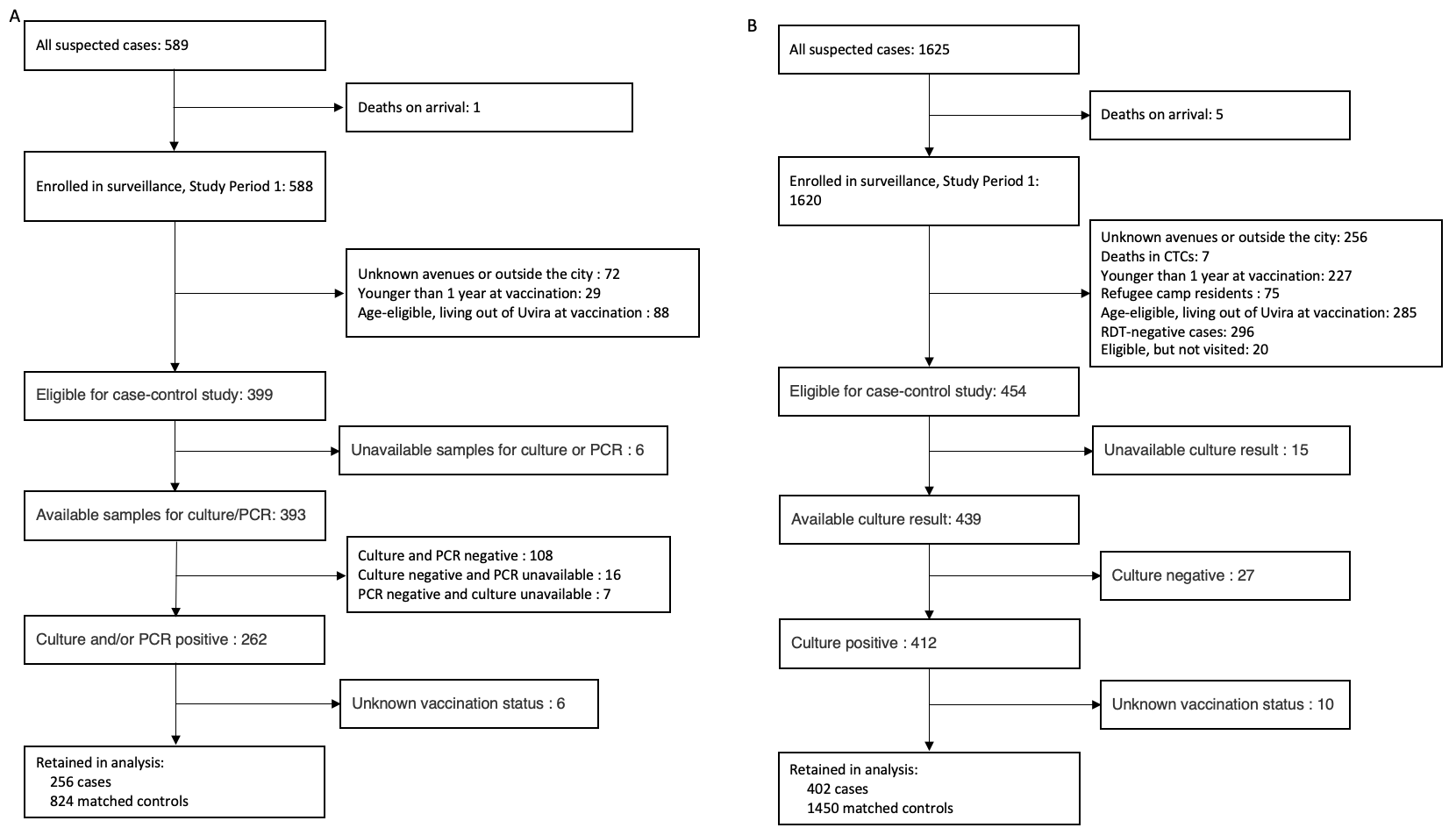

**Figure S1. Flow chart of recruitment of participants by study period.** On left (A), the enrolment process for participants in the analysis of the cholera vaccine effectiveness 12-17 months after vaccination (SP1), and on right (B) participants in the 24-36 months post-vaccination (SP2) analysis. Cases with unavailable culture results are those for which suspected colonies were isolated and oxidase test was positive at the onsite laboratory, but that did not have agglutination results because of a stockout of antiserum or attempts to regrow them at the reference laboratory in Goma were unsuccessful.

#### 2.2. Description of participants

**Table S2. Description of cases by Study Period**

| Characteristic | Overall,  N = 658 | Study Period 1,  N = 256 | Study Period 2,  N = 402 | P value |
| --- | --- | --- | --- | --- |
| Age (years), median (IQR)* | 14.1 (6.0, 33.8) | 16.8 (7.6, 35.8) | 11.5 (5.9, 30.0) | 0.005 |
| Age group (years) |  |  |  | 0.006 |
| 1-4 | 99 (15.0%) | 39 (15.2%) | 60 (14.9%) |  |
| 5-9 | 146 (22.2%) | 37 (14.5%) | 109 (27.1%) |  |
| 10-19 | 159 (24.2%) | 69 (27.0%) | 90 (22.4%) |  |
| 20-39 | 124 (18.8%) | 57 (22.3%) | 67 (16.7%) |  |
| 40-59 | 91 (13.8%) | 36 (14.1%) | 55 (13.7%) |  |
| ≥60 | 39 (5.9%) | 18 (7.0%) | 21 (5.2%) |  |
| Sex |  |  |  | 0.145 |
| Female | 337 (51.2%) | 122 (47.7%) | 215 (53.5%) |  |
| Male | 321 (48.8%) | 134 (52.3%) | 187 (46.5%) |  |
| Health facility |  |  |  | 0.715 |
| CTC (Uvira referral hospital) | 572 (86.9%) | 221 (86.3%) | 351 (87.3%) |  |
| CTU (Kalundu CEPAC health center) | 86 (13.1%) | 35 (13.7%) | 51 (12.7%) |  |
| Level of dehydration** |  |  |  | 0.104 |
| Mild | 12 (1.8%) | 6 (2.3%) | 6 (1.5%) |  |
| Moderate | 241 (36.6%) | 105 (41.0%) | 136 (33.8%) |  |
| Severe | 405 (61.6%) | 145 (56.6%) | 260 (64.7%) |  |
| Hospitalization duration(days) |  |  |  | 0.071 |
| Zero | 11 (1.7%) | 7 (2.7%) | 4 (1.0%) |  |
| 1-2 | 295 (44.8%) | 104 (40.6%) | 191 (47.5%) |  |
| ≥ 3 | 352 (53.5%) | 145 (56.6%) | 207 (51.5%) |  |
| Time from symptoms onset to hospitalization (days) |  |  |  | 0.214 |
| Zero | 390 (59.3%) | 156 (60.9%) | 234 (58.2%) |  |
| 1 | 263 (40.0%) | 100 (39.1%) | 163 (40.5%) |  |
| ≥2 | 5 (0.8%) | 0 (0.0%) | 5 (1.2%) |  |
| Treated at another health facility before admission to CTC/CTU | 112 (17.0%) | 38 (14.8%) | 74 (18.4%) | 0.236 |
| Treated at a pharmacy before admission | 239 (36.3%) | 45 (17.6%) | 194 (48.3%) | <0.001 |
| Treated by traditional healer before admission | 2 (0.3%) | 1 (0.4%) | 1 (0.2%) | >0.999 |
| Used antibiotics before admission |  |  |  | <0.001 |
| Yes | 345 (69.0%) | 139 (73.2%) | 206 (66.5%) |  |
| No | 117 (23.4%) | 51 (26.8%) | 66 (21.3%) |  |
| Uncertain (used unspecified drugs)*** | 38 (7.6%) | 0 (0.0%) | 38 (12.3%) |  |
| Missing | 158 | 66 | 92 |  |
| Admitted to CTC/CTU before for diarrhea | 28 (4.3%) | 9 (3.6%) | 19 (4.7%) | 0.474 |
| Missing | 5 | 4 | 1 |  |
| Has been told she/he had cholera before | 28 (4.3%) | 8 (3.2%) | 20 (5.0%) | 0.256 |
| Missing | 6 | 3 | 3 |  |
| Overall vaccination status |  |  |  | 0.938 |
| Not Vaccinated | 452 (68.7%) | 177 (69.1%) | 275 (68.4%) |  |
| One Dose | 133 (20.2%) | 52 (20.3%) | 81 (20.1%) |  |
| Two Doses | 73 (11.1%) | 27 (10.5%) | 46 (11.4%) |  |
| Vaccination status (≥ 1 dose) |  |  |  | 0.843 |
| Not vaccinated | 452 (68.7%) | 177 (69.1%) | 275 (68.4%) |  |
| One dose or more | 206 (31.3%) | 79 (30.9%) | 127 (31.6%) |  |
| Vaccination status (single dose) |  |  |  | 0.990 |
| Not vaccinated | 452 (77.3%) | 177 (77.3%) | 275 (77.2%) |  |
| Single dose | 133 (22.7%) | 52 (22.7%) | 81 (22.8%) |  |
| Vaccination card available | 29 (14.1%) | 7 (8.9%) | 22 (17.3%) | 0.090 |

The characteristics of study cases were compared using the Wilcoxon rank sum and Pearson's Chi-squared (or Fisher's exact) tests. *Age refers to the age on the first day of the second mass vaccination campaign round (01 October 2020).

**The level of dehydration assessed using the Global Task Force on Cholera Control (GTFCC) guidance.^4^ ***Uncertain means that the patient could not recall the name or type of the medicines they took before admission to the cholera treatment facility.

**Table S3. Characteristics of participants in the 12-17 months vaccine effectiveness study**

| Characteristic | Overall,  N = 1,080 | Cases,  N = 256 | Controls,  N = 824 | P value** | SMD |
| --- | --- | --- | --- | --- | --- |
| Age group at vaccination (years)* |  |  |  |  | 0.075 |
| 1-4 | 148 (13.7%) | 39 (15.2%) | 109 (13.2%) | Reference |  |
| 5-9 | 151 (14.0%) | 37 (14.5%) | 114 (13.8%) | 0.0524 |  |
| 10-19 | 298 (27.6%) | 69 (27.0%) | 229 (27.8%) | 0.0242 |  |
| 20-39 | 256 (23.7%) | 57 (22.3%) | 199 (24.2%) | 0.0252 |  |
| 40-59 | 156 (14.4%) | 36 (14.1%) | 120 (14.6%) | 0.0093 |  |
| ≥60 | 71 (6.6%) | 18 (7.0%) | 53 (6.4%) | 0.0130 |  |
| Sex |  |  |  |  | 0.003 |
| Female | 516 (47.8%) | 122 (47.7%) | 394 (47.8%) | Reference |  |
| Male | 564 (52.2%) | 134 (52.3%) | 430 (52.2%) | 0.7154 |  |
| Vaccination status |  |  |  |  | 0.356 |
| Not Vaccinated | 607 (56.2%) | 177 (69.1%) | 430 (52.2%) | Reference |  |
| One Dose | 326 (30.2%) | 52 (20.3%) | 274 (33.3%) | <0.0001 |  |
| Two Doses | 147 (13.6%) | 27 (10.5%) | 120 (14.6%) | 0.0044 |  |
| Vaccination card available | 61 (12.9%) | 7 (8.9%) | 54 (13.7%) | 0.9618 | 0.153 |

*Age refers to the age on the first day of the second mass vaccination campaign round (01 October 2020) and is reported as median (interquartile range). SMD: Standardized mean difference. ** P values come from univariate conditional logistic regression models.

**Table S4. Univariate conditional logistic regression model of factors associated with cholera in Study Period 2**

| Variable | OR (95% CI) | P value |
| --- | --- | --- |
| Age at vaccination (years) | 0.99 (0.96–1.02) | 0.3896 |
| Age group at vaccination (years) |  |  |
| 1-4 | Reference |  |
| 5-9 | 1.59 (0.99–2.57) | 0.0563 |
| 10-19 | 0.7 (0.35–1.38) | 0.3003 |
| 20-39 | 0.43 (0.17–1.09) | 0.0738 |
| 40-59 | 0.61 (0.18–2.07) | 0.4303 |
| ≥60 | 0.54 (0.11–2.6) | 0.4427 |
| Sex |  |  |
| Female | Reference |  |
| Male | 0.96 (0.28–3.29) | 0.9500 |
| Level of education |  |  |
| None or primary | Reference |  |
| Lower secondary | 0.49 (0.26–0.92) | 0.0277 |
| Upper secondary | 0.39 (0.22–0.67) | 0.0008 |
| Bachelor or higher | 0.63 (0.23–1.67) | 0.3492 |
| Occupation |  |  |
| None | Reference |  |
| Preschool children | 0.82 (0.40–1.66) | 0.5777 |
| Students | 0.9 (0.52–1.57) | 0.7115 |
| Informal work | 0.75 (0.51–1.11) | 0.1483 |
| Salaried | 0.48 (0.24–0.98) | 0.0425 |
| Household size | 1.02 (0.99–1.06) | 0.2162 |
| Living in household with unimproved drinking water source | 1.12 (0.83–1.52) | 0.4573 |
| Living in household with shared toilet | 1.41 (1.11–1.79) | 0.0049 |
| Living in household with unimproved toilet | 1.24 (0.95–1.61) | 0.1117 |
| Availability of soap and water for handwashing | 1.86 (1.40–2.48) | <0.0001 |
| Living in house with electricity | 0.72 (0.56–0.92) | 0.0090 |
| Wealth index | 0.58 (0.44–0.76) | 0.0001 |
| Vaccination status |  |  |
| Not vaccinated | Reference |  |
| One dose | 0.53 (0.4–0.71) | <0.0001 |
| Two doses | 0.85 (0.58–1.24) | 0.3868 |
| Vaccination card available* | 2.17 (1.1–4.27) | 0.0252 |

*: Estimates for the availability of vaccination card are based only on people who receipt of at least one dose of the cholera vaccine.

##### 2.3. Oral cholera vaccine effectiveness sensitivity analyses

##### 2.3.1. Single-dose vaccine effectiveness by possession of a vaccination card

There were very few participants able to produce their vaccination cards when asked. And there is no cholera vaccination register in Uvira, making self-reporting the only source of information about vaccination status for most people. When we restricted the VE analysis only to study participants with vaccination status confirmed by cards (and those who report being unvaccinated), our overall point estimates remained comparable to estimates from analysis including those whose vaccination cards were unavailable, though with very large confidence intervals, as expected (Table S5).

**Table S5. Single-dose vaccine effectiveness estimates for the entire study period when considering only those who reported a vaccination card as vaccinated and considering those who reported to be vaccinated without a card as missing data.**

| Population | Cases (Effective N) | Controls (Effective N) | Unadjusted VE (95% CI) | Adjusted VE (95% CI) |
| --- | --- | --- | --- | --- |
| **12-36 months after vaccination** | | | | |
| Overall | 573 (419) | 1998 (763) | 47.8% (34.6–58.4) | 48.1% (34.6–58.7) |
| 1-4 years | 96 (61) | 263 (129) | 52.4% (22.5–70.8) | 48.7% (13.2–69.6) |
| ≥5 years | 463 (332) | 1534 (584) | 46.7% (31.8–58.4) | 47.9% (32.9–59.5) |
| Vaccinated participants showing a vaccination card | | | | |
| Overall | 427 (53) | 1019 (79) | 46.1% (-7.3–73) | 47.8% (-4.5–74.0) |
| 1-4 years | 61 (4) | 106 (9) | 70.6% (-159.3–96.7) | 65.9% (-202.2–96.1) |
| ≥5 years | 350 (45) | 836 (66) | 38.8% (-22.4–69.4) | 43.0% (-15.2–71.7) |

##### 2.3.2 Alternative regression models for the period 24-36 months post-vaccination

We explore the robustness of our VE estimates by fitting 4 models with different sets of covariates for the Study Period 2 (Table S6). The final model includes age, household size, household wealth index, type of sanitation facility, whether the participant used a toilet shared by multiple households compared to using a private toilet, drinking water sources, and availability of a hand washing facility and soap. Model variant 1 is a slightly different version of the final model, with the variable about whether the participant lives in a house with electricity replacing the wealth index. In Model variant 2 we only adjusted for the WASH variables and age, while Model variant 3 accounts for all the variables (including all WASH variables) that had an SMD ≥ 0.1 in the bivariable comparisons (Table 2).

**Table S6. Regression models variants for Study Period 2 (24-36 months after vaccination)**

|  | **Adjusted VE (95% CI)** | **AIC** | **Confounders** |
| --- | --- | --- | --- |
| **Final model** |  | 950.3 |  |
| Overall | 45.5 (25.8–60.0) |  | age_vacc, n_household, occup, wealth_index, toilet_shared, drinking_water_source, handwash_soapwater |
| 1-4 years | 33.1 (-30–65.6) |  |  |
| ≥5 years | 48.3 (27.2–63.3) |  |  |
| **Model variant 1** |  | 957.8 |  |
| Overall | 45.7 (26.2–60.0) |  | age_vacc, n_household, occup, electricity, toilet_shared, drinking_water_source, handwash_soapwater |
| 1-4 years | 31.9 (-32.2–64.9) |  |  |
| ≥5 years | 48.8 (28–63.5) |  |  |
| **Model variant 2** |  | 966.8 |  |
| Overall | 45.2 (26.1–59.4) |  | age_vacc, toilet_shared, drinking_water_source, handwash_soapwater |
| 1-4 years | 33.5 (-27.3–65.3) |  |  |
| ≥5 years | 47.9 (27.4–62.6) |  |  |
| **Model variant 3** |  | 961.1 |  |
| Overall | 45.5 (26.1–59.8) |  | age_vacc, occup, electricity, wealth_index, toilet_shared, drinking_water_source, handwash_soapwater |
| 1-4 years | 34.1 (-27.8–66) |  |  |
| ≥5 years | 48.1 (27.2–63.0) |  |  |

*age_vacc*: age (in years) at the time of vaccine, *n_household*: household size, *ind_school_ever*: whether a person has ever gone to school, *occup*: occupation, *electricity:* electricity in household, *wealth_quintile*: wealth quintile, *toilet_shared*: whether the participant lives in a household that shares a toilet with other households, *toilet_type*: whether the participant’s household use an improved sanitation facility; *drinking_water_source*: whether the participant’s household has used unimproved drinking water source in the week prior to interview; *handwash_soap*: whether a handwashing facility and soap were available at the time of visit.

In Study Period 2, three continuous covariates were included in VE regression models. We explored models with these covariates as linear terms in addition to polynomials (of two and three degrees) and restricted cubic splines (with four knots). We then compared those models with a base model with linear effects. The model with quadratic term for household size was the best combination used for main single-dose VE estimates (Table S7).

**Table S7. Regression models with different functional forms for continuous covariates.**

| Continuous covariate with non-linear effect | Adjusted VE (95% CI) | AIC | P-value, Likelihood ratio test |
| --- | --- | --- | --- |
| Model with linear functions for all continuous covariates |  |  |  |
| Overall | 45.1 (25.7–59.4) | 966.2 |  |
| 1-4 years | 41.4 (-11.9–69.3) |  |  |
| ≥5 years | 46 (24.5–61.4) |  |  |
| Best model* |  |  |  |
| Overall | 45.5 (25.8–60) | 950.3 | 0.0001 |
| 1-4 years | 33.1 (-30–65.6) |  |  |
| ≥5 years | 48.3 (27.2–63.3) |  |  |
| Age, quadratic |  |  |  |
| Overall | 45.3 (25.8–59.7) | 959.6 | 0.0035 |
| 1-4 years | 32.6 (-31.1–65.4) |  |  |
| ≥5 years | 48.2 (27.3–63) |  |  |
| Age, 3-degree polynomial |  |  |  |
| Overall | 45.4 (25.9–59.8) | 961.4 | 0.0123 |
| 1-4 years | 31.8 (-33–65.1) |  |  |
| ≥5 years | 48.5 (27.7–63.3) |  |  |
| Age, RCS |  |  |  |
| Overall | 45.5 (26.1–59.8) | 960.8 | 0.0090 |
| 1-4 years | 35.1 (-27–66.8) |  |  |
| ≥5 years | 47.9 (26.9–62.9) |  |  |
| Household size, quadratic |  |  |  |
| Overall | 45.8 (26.5–60) | 960.0 | 0.0041 |
| 1-4 years | 41.2 (-12.2–69.2) |  |  |
| ≥5 years | 46.9 (25.6–62.1) |  |  |
| Household size, 3-degree polynomial |  |  |  |
| Overall | 45.3 (25.7–59.7) | 955.4 | 0.0006 |
| 1-4 years | 41.2 (-12.4–69.2) |  |  |
| ≥5 years | 46.3 (24.6–61.8) |  |  |
| Household size, RCS |  |  |  |
| Overall | 44.8 (25.2–59.2) | 965.7 | 0.1046 |
| 1-4 years | 39.7 (-15.2–68.5) |  |  |
| ≥5 years | 46 (24.4–61.4) |  |  |
| Wealth index, quadratic |  |  |  |
| Overall | 45.1 (25.7–59.4) | 968.1 | 0.8235 |
| 1-4 years | 41.5 (-11.8–69.4) |  |  |
| ≥5 years | 45.9 (24.4–61.3) |  |  |
| Wealth index, 3-degree polynomial |  |  |  |
| Overall | 45 (25.6–59.4) | 970.1 | 0.9648 |
| 1-4 years | 41.4 (-11.9–69.3) |  |  |
| ≥5 years | 45.9 (24.4–61.3) |  |  |
| Wealth index, RCS |  |  |  |
| Overall | 45 (25.6–59.3) | 969.9 | 0.8758 |
| 1-4 years | 41.4 (-11.8–69.3) |  |  |
| ≥5 years | 45.9 (24.3–61.3) |  |  |
| Age, household size and wealth index, all quadratic |  |  |  |
| Overall | 46 (26.6–60.2) | 955.4 | 0.0008 |
| 1-4 years | 32.3 (-31.4–65.2) |  |  |
| ≥5 years | 49 (28.3–63.7) |  |  |
| Age, household size and wealth index, all 3-degree polynomial |  |  |  |
| Overall | 45.6 (25.9–60.1) | 955.5 | 0.0009 |
| 1-4 years | 31.7 (-33.1–65) |  |  |
| ≥5 years | 48.7 (27.7–63.6) |  |  |
| Age, household size and wealth index, all RCS |  |  |  |
| Overall | 45 (25.3–59.5) | 964.7 | 0.0364 |
| 1-4 years | 34 (-29–66.2) |  |  |
| ≥5 years | 47.6 (26.2–62.7) |  |  |

RCS: restricted cubic splines. *The best model incorporates a quadratic function for age and a 3-degree polynomial function for household size.

#### 2.3.3. Effectiveness of at least one dose of oral cholera vaccine

In secondary analysis, we estimated for receipt of at least 1 dose, comparing individuals who reported having received one or more doses of OCV to those who did not receive any dose (Table S8).

**Table S8. Effectiveness of at least one dose of oral cholera vaccine, 12-17 months and 24-36 months after vaccination campaigns**

| Population | Cases (effective n) | Controls (effective n) | Unadjusted VE (95% CI) | Adjusted VE (95% CI)* |
| --- | --- | --- | --- | --- |
| **12-36 months after vaccination** | | | | |
| Overall | 644 (519) | 2273 (988) | 45.3% (33.3–55.1) | 45.6% (33.5–55.5) |
| 1-4 years | 120 (79) | 295 (157) | 38.8% (9.6–58.5) | 30.7% (-6.2–54.8) |
| ≥5 years | 506 (405) | 1741 (772) | 46.7% (33.8–57.1) | 48.6% (35.8–58.9) |
| **12-17 months after vaccination (Study Period 1)** | | | | |
| Overall | 245 (208) | 823 (399) | 53.3% (36.1–65.9) | 53.1% (35.2–66.0) |
| 1-4 years | 43 (30) | 94 (59) | 56.3% (12.1–78.3) | 63.8% (21.8–83.3) |
| ≥5 years | 194 (168) | 663 (323) | 52.7% (33.7–66.3) | 50.6% (29.7–65.3) |
| **24-36 months after vaccination (Study Period 2)** | | | | |
| Overall | 399 (311) | 1450 (589) | 39.1% (21.3–52.8) | 40.0% (21.4–54.1) |
| 1-4 years | 77 (49) | 201 (98) | 27% (-17.4–54.6) | -0.9% (-76.6–42.3) |
| ≥5 years | 312 (237) | 1078 (449) | 41.8% (22.9–56.1) | 47.5% (29.3–61.0) |

Effective n represents the number of cases or controls in case-control sets with non-identical vaccination status. *: In SP1 and in analyses combining data from both study periods, we only adjusted for age as a continuous variable.

#### 2.3.4. Effectiveness of two doses of oral cholera vaccine

We also examined the VE of two-doses, considering those reporting receipt of a single dose as missing. The unadjusted and adjusted VE (95% CI) for two doses was 56·1% (95% 24·8–74·4) and 57·1% (24·9–75·5)) in Study Period 1 for individuals aged five years and older. The small effective sample size of children 1-4 years old in SP1 and in all age groups in SP 2 challenges the interpretation of the point estimates of VE in these strata (Table S9). As shown in the Table S10 below, we had extremely low power for these estimates.

**Table S9. Effectiveness of two doses of oral cholera vaccine, 12-17 months and 24-36 months after vaccination campaigns.**

| Population | Cases (effective n) | Controls (effective n) | Unadjusted VE (95% CI) | Adjusted VE (95% CI) |
| --- | --- | --- | --- | --- |
| **12-36 months after vaccination*** | | | | |
| Overall | 513 (197) | 1556 (342) | 40.0% (15.9–57.2) | 40.2% (15.7–57.6) |
| 1-4 years | 93 (27) | 186 (50) | -19.1% (-125.9–37.2) | -41.8% (-176.1–27.2) |
| ≥5 years | 407 (151) | 1227 (259) | 49.0% (25.4–65.2) | 51.3% (28.2–67) |
| **12-17 months after vaccination (Study Period 1)*** | | | | |
| Overall | 194 (88) | 550 (141) | 56.1% (24.8–74.4) | 57.9% (26.5–75.9) |
| 1-4 years | 34 (11) | 58 (21) | 14.9% (-145–70.4) | 19.4% (-149.7–74) |
| ≥5 years | 154 (70) | 453 (110) | 61.6% (29.7–79.1) | 63.1% (30.7–80.4) |
| **24-36 months after vaccination (Study Period 2)** | | | | |
| Overall | 319 (109) | 1006 (201) | 24.6% (-16.7–51.3) | 25.4% (-17.8–52.7) |
| 1-4 years | 59 (16) | 128 (29) | -48.4% (-233.6–34) | -148.2% (-500.4–-2.6) |
| ≥5 years | 253 (81) | 774 (149) | 36.7% (-3.5–61.3) | 45.3% (8.4–67.4) |

*: In SP1 and in analyses combining data from both study periods, we only adjusted for age as a continuous variable, in quadratic form.

#### 2.3.8. Effective study power

We explored the effective study power based on the number cases and controls that contributed to the VE effectiveness estimates, that is, the number of study participants in matched case-controls sets where at least one control had a vaccination status different from that of the case. We assumed a vaccine effectiveness of 50% for individuals aged at least 5 years (in analysis including all ages), and 37·5% for individuals 1-4 years old (that is, 25% lower than in older individuals). We used the *epi.sscc* function of the *epiR* package in R for power calculations. For each age stratum, we derived the proportions of vaccinated controls from the study sample.

The study power for the 1-4-year age group was weak across all analyses, explaining the uncertainty around VE estimates in this age group, even when combining data from both study periods. The study power was also weak in Study Period 2, far weaker for two-dose VE analysis than for single-dose analysis. We only had a study power 14·2% to detect a significant two-dose VE in all age groups in Study Period 2 (Table S10).

**Table S10. Effective study power for one and two dose estimates of effectiveness**

| Population | Total sample | Cases | Controls | Odds ratio | Power |
| --- | --- | --- | --- | --- | --- |
| **Single-dose VE analysis, 12-36 months since vaccination** | | | | | |
| Overall | 849 | 425 | 424 | 0.500 | 0.993 |
| 1-4 years | 171 | 55 | 116 | 0.625 | 0.240 |
| ≥5 years | 640 | 320 | 320 | 0.500 | 0.965 |
| **Two-dose VE analysis, 12-36 months since vaccination** | | | | | |
| Overall | 415 | 208 | 207 | 0.500 | 0.509 |
| 1-4 years | 72 | 25 | 47 | 0.625 | 0.051 |
| ≥5 years | 317 | 159 | 158 | 0.500 | 0.415 |
| **Single-dose VE analysis, 12-17 months since vaccination** | | | | | |
| Overall | 516 | 184 | 332 | 0.500 | 0.783 |
| 1-4 years | 82 | 27 | 55 | 0.625 | 0.137 |
| ≥5 years | 417 | 150 | 267 | 0.500 | 0.686 |
| **Two-dose VE analysis, 12-17 months since vaccination** | | | | | |
| Overall | 254 | 97 | 157 | 0.500 | 0.300 |
| 1-4 years | 44 | 16 | 28 | 0.625 | 0.044 |
| ≥5 years | 202 | 79 | 123 | 0.500 | 0.252 |
| **Single-dose VE analysis, 24-36 months since vaccination** | | | | | |
| Overall | 333 | 114 | 219 | 0.500 | 0.525 |
| 1-4 years | 89 | 28 | 61 | 0.625 | 0.135 |
| ≥5 years | 223 | 77 | 146 | 0.500 | 0.359 |
| **Two-dose VE analysis, 24-36 months since vaccination** | | | | | |
| Overall | 161 | 51 | 110 | 0.500 | 0.142 |
| 1-4 years | 28 | 9 | 19 | 0.625 | 0.034 |
| ≥5 years | 115 | 38 | 77 | 0.500 | 0.111 |

#### 2.3.6. Potential misclassification of vaccination status in two-dose estimates

Field workers and clinicians involved in the study raised the possibility that some unvaccinated people would report being fully vaccinated, rather than partially, due to social desirability bias and the fact that when asking about whether each person was vaccinated, the study interviewer tells the participants that it is a two-dose vaccine. Given that our point estimates for two-dose effectiveness were lower than expected (though with very wide confidence intervals), we conducted a simple simulation analysis to understand how many cases that reported having had two doses would need to be misclassified to have a point estimate consistent with the one dose VE estimates. In these simulations we found that if 8-9 cases who reported having had two doses were truly unvaccinated, our two dose point estimates would reach those of one dose VE presented in the main analysis (Figure S2).

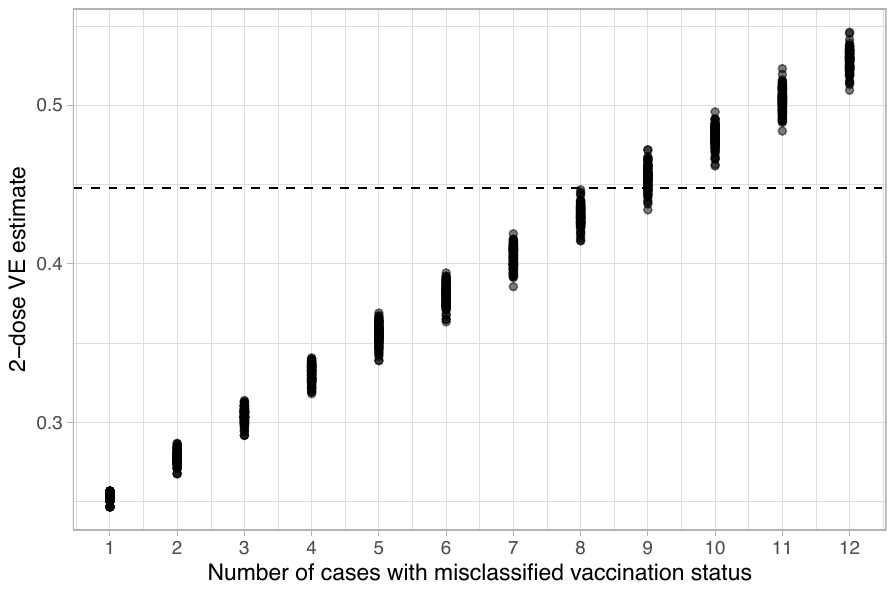

**Figure S2. Estimated number of cases with misclassification of vaccination status necessary to lead to VE estimates comparable to that of a single dose.**

#### 2.3.5. Effectiveness of one dose of oral cholera vaccine using culture as confirmation test in Study Period 1

In Table S11 we present kOCV effectiveness estimates from sensitivity analysis using culture alone for cholera confirmation in Study Period 1, without considering PCR testing, similarly to Study Period 2

**Table S11. Effectiveness of single dose of oral cholera vaccine, using culture alone for cholera confirmation. Note that SP2 only used culture, so results are the same as in the main text.**

| Population | Cases (Effective N) | Controls (Effective N) | Unadjusted VE (95% CI) | Adjusted VE (95% CI) |
| --- | --- | --- | --- | --- |
| **Duration: 12-36 months** | | | | |
| Overall | 482 (349) | 1712 (732) | 50.1% (35.9–61.2) | 50.3% (36.0–61.4) |
| 1-4 years | 78 (49) | 223 (119) | 52.7% (19.4–72.2) | 44.2% (1.3–68.5) |
| ≥5 years | 391 (276) | 1309 (560) | 49.5% (33.6–61.6) | 51.6% (36.1–63.4) |
| **Duration: 12-17 months** | | | | |
| Overall | 128 (100) | 418 (269) | 65.5% (42.9–79.2) | 64.2% (40.4–78.5) |
| 1-4 years | 16 (11) | 43 (34) | 80.8% (12.2–95.8) | 80.7% (9.6–95.9) |
| ≥5 years | 107 (85) | 343 (219) | 61.4% (35.2–77) | 59.7% (31.7–76.2) |

#### 2.2. Description of participants

| Round | N | Months after 2nd dose campaign | At least one dose (95% CI) | At least two doses (95% CI) |
| --- | --- | --- | --- | --- |
| Survey 1 | 2,292 | 11.0 - 11.7 | 55% (51-60) | 23% (20-27) |
| Survey 2 | 3,583 | 19.5 - 20.6 | 47% (44-50) | 20% (18-23) |
| Survey 3 | 2,864 | 32.5 - 33.2 | 39% (36-43) | 10% (8-12) |

### References

1	Jeandron A, Cumming O, Rumedeka BB, Saidi JM, Cousens S. Confirmation of cholera by rapid diagnostic test amongst patients admitted to the cholera treatment centre in Uvira, Democratic Republic of the Congo. *PLoS One* 2018; **13**: e0201306.

2	The WHO/UNICEF Joint Monitoring Programme for Water Supply, Sanitation and Hygiene (JMP). JMP Methodology 2017 Update & SDG Baselines. WHO/UNICEF JMP, 2018 <https://washdata.org/report/jmp-methodology-2017-update> (accessed July 9, 2023).

3	Filmer D, Pritchett LH. Estimating wealth effects without expenditure data-or tears: An application to educational enrollments in states of India. *Demography* 2001; **38**: 115.

4	Global Task Force on Cholera Control (Surveillance Working Group). Interim guidance document on cholera surveillance. GTFCC, 2017 <https://www.gtfcc.org/wp-content/uploads/2019/10/gtfcc-interim-guidance-document-on-cholera-surveillance.pdf> (accessed Aug 3, 2023).
